# SEIR Transmission dynamics model of 2019 nCoV coronavirus with considering the weak infectious ability and changes in latency duration

**DOI:** 10.1101/2020.02.16.20023655

**Authors:** Shi Pengpeng, Cao Shengli, Feng Peihua

**Affiliations:** School of Civil Engineering & Institute of Mechanics and Technology, Xi’an University of Architecture and Technology, Xi’an 710055, Shaanxi, China; School of Energy and Power Engineering, Xi’an Jiaotong University, Xi’an 710049, Shaanxi, China; State Key Laboratory for Strength and Vibration of Mechanical Structures, Xi’an Jiaotong University, Xi’an 710049, Shaanxi, China

## Abstract

Pneumonia patients of 2019-ncov in latent period are not easy to be effectively quarantined, but there is evidence that they have strong infectious ability. Here, the infectious ability of patients during the latent period is slightly less than that of the infected patients was assumed. We established a new SEIR propagation dynamics model, that considered the weak transmission ability of the incubation period, the variation of the incubation period length, and the government intervention measures to track and isolate comprehensively. Based on the raw epidemic data of China from January 23, 2020 to February 10, 2020, the dynamic parameters of the new present SEIR model are fitted. Through the Euler integration algorithm to solve the model, the effect of infectious ability of incubation patients on the theoretical estimation of the present SEIR model was analyzed, and the occurrence time of peak number in China was predicted.

---

The new coronavirus (2019-nCoV) is a major epidemic that humans are experiencing. As a kind of coronavirus, it poses a continuing threat to human health because of its high transmission efficiency and serious infection consequences ^**[1]**^. Since the first case of novel coronavirus pneumonia was discovered in early Dec 2019, it has spread widely over the past two months. By the end of 10:00, Feb 10th, 2020, there were 42708 confirmed cases of 2019-nCoV infection and 21675 suspected cases in China. At present, 2019-nCoV pneumonia cases have been confirmed in dozens of countries and regions including the United States, Germany, France, Canada, Australia, and Japan.

The rapid spread of 2019-noV pneumonia around the world poses a major threat to the closely connected and interdependent world today. The reproductive number R refers to the average number of secondary cases generated from primary cases, and has become a key quantity to determine the intensity of interventions needed to control epidemics^**[2]**^. On January 29, 2020, Li et al conducted a study of the first 425 confirmed cases in Wuhan, showing that the R of 2019-nCoV was 2.2, and revealed that person-to-person transmission occurred between close contacts ^**[3]**^. On January 26, 2020, new research shows that the reproductive ratio of 2019-nCoV is 2.90, which is higher than the 1.77 SARS epidemic^**[4]**^° In the absence of comprehensive treatments or vaccines, China has adopted the most effective isolation prevention and control measures to isolate patients diagnosed with 2019-nCoV to control the spread of infection, but the number of 2019-nCoV infections has far exceeded that of SARS epidemics. Existing basic research results and actual situation of epidemic spread have shown that 2019-nCoV has a higher pandemic risk than the outbreak of SARS in 2003^**[4]**^°

This paper discusses the feasibility of using the deterministic model of the transmission dynamics model of infectious diseases to assess the development of the 2019-nCoV epidemic in China. Some researchers have tried to study and evaluate the development trend of the 2019-nCoV pneumonia epidemic through the transmission dynamics. On Jan 24, 2020, British scholars Read et al^**[5]**^ estimated that the number of 2019-nCoV infection cases in Wuhan would reach 190000 on Feb 4, which obviously overestimated the development trend of the epidemic. On Jan 31, 2020, Wu et al^**[6]**^, scholars from Hong Kong, China, predicted that the number of infections in Wuhan on Jan 25 exceeded 75815, which also obviously overestimated the spread of the epidemic. Chinese scholars Shen et al. ^**[7]**^ used dynamics models to predict the peak time and scale of the epidemic, and estimated that the peak number of infections was less than 20,000, which was lower than the raw data released on Feb 10. Chinese scholars Xiao et al. ^**[8]**^ established a transmission dynamic model with considering intervention strategies such as close tracking and quarantine. Based on their model, scholars predicted that the epidemic would reach its peak around Feb 5^**[8]**^, which made an early estimation of the peak time of the epidemic. In summary, although some researchers have carried out research on the transmission dynamics of the 2019-nCoV pneumonia epidemic, the real epidemic development has far deviated from the prediction results of previous studies.

In previous studies on the transmission dynamics of infectious diseases, the epidemic was in the early stages of development and lacked sufficient raw data, so it was difficult to accurately predict the development of the epidemic. In addition, the shortcomings of the transmission dynamic model itself have also become the direct cause of limiting its prediction effect. Latent patients are not easy to be effectively quarantined, and recent evidence shows that latent patients have a strong infectious ability, but the existing epidemic transmission dynamic models ^**[5-8]**^ often ignore the transmission risks caused by patients in the latent period. In addition, researchers have found that estimates of the average latency of 2019-nCoV are also changing. The incubation period was determined to be 7 days in January, and it was recently estimated to be 3 days, which means that 2019-nCoV infected people become more likely to develop symptoms.

Obviously, it is still necessary to make a new assessment of the spread of the 2019-nCoV epidemic situation, which has important practical significance for the research, judgment and prevention of the epidemic situation. In this paper, we established a new SEIR propagation dynamics model, considering the weak transmission ability of the incubation period, the variation of the incubation period length, and the government intervention measures to track and quarantine comprehensively. Based on this new SEIR propagation dynamics model, the effect of infectious ability of incubation patients on the theoretical estimation of the present SEIR model was analyzed, and the occurrence time of peak number in China was predicted.

Considering that the Chinese Spring Festival began on January 23 and the Chinese government initiated effective prevention, control and quarantine measures for the whole people, it can be considered that there are few new imported cases in all provinces, and the impact of migrant population flow can be basically ignored. Therefore, the premise of carrying out dynamic model research on infectious disease transmission is sufficient. The model in this paper is based on the classical SEIR model. The classic SEIR model divides the population into *S* (Susceptible), *I* (Infected), *E* (Exposed), and *R* (Recovered). In this model, it is also assumed that all individuals in the population are at risk of infection. Antibodies will be produced when the infected individual recovers, so the recovered population *R* will not be infected again. In addition, quarantine measures to prevent and treat infectious diseases are also taken into account in the model. The population components in the model also includes *Sq* (isolated susceptible), isolation exposed *Eq* (isolated exposed) and *Iq* (isolated infected). In view of the fact that isolated infected individuals will be immediately sent to designated hospitals for quarantine and treatment, this part of the population will be converted to hospitalized *H* in this model. Therefore, the original populations *S, I*, and *E* respectively refer to susceptible, infected, and exposed individuals who have missed quarantine measures. The isolated susceptible person is re-converted to the susceptible person after isolation. Both the infected and the exposed have the ability to infect the susceptible, turning them into the exposed. The transformation relationship among different groups of people is shown in Fig. 1.

**Figure 1.**
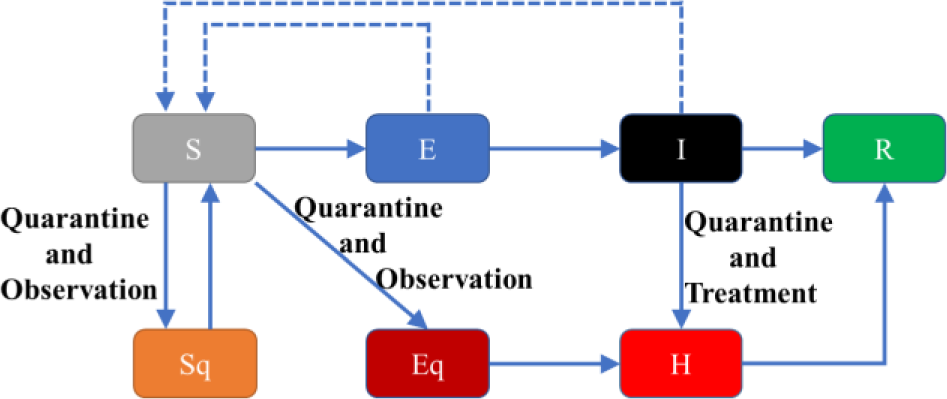
Modified SEIR propagation dynamics model.

It is assumed that the quarantine ratio is *q*, the probability of transmission is *β*, and the contact rate is *c*. The transformation rates of *S* to *Sq, Eq* and *E* are *cq*(1 − *β*), *cβ q* and *cβ* (1 − *q*), respectively. Therefore, the control equation for the number of susceptible persons is

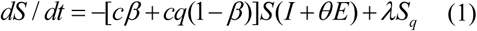

where *θ* is the ratio of the transmission ability of the exposed to the infected. When *θ* =0, it means that the infection ability of the patients in the latent period is ignored. When *θ* =1, the infection ability of the patients in the latent period is the same as that of the patients with symptoms. And *λ* is the rate of isolation release, hence *λ* = 1/ 14(the quarantine duration is 14 days).

With the growth of statistical data, researchers’ estimation of the average latent period of 2019-nCoV is also changing. The average incubation period has changed from 7 days determined in January to 3 days estimated recently, which means 2019-ncov infected people are more likely to show symptoms. *σ* is the transformation rate from the exposed to the infected. Considering the change of actual latent period, a smooth transition curve from 1/7 to 1/3 is used to represent the change law of transformation rate from the exposed to infected under the change of the latent period. The curve equation is

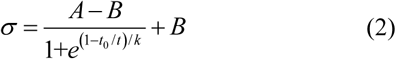

where *A* and *B* correspond to the values of the transformation rate of the exposed to the infected in the early and late stages of the epidemic period studied. The epidemic data in this letter are from Jan. 23 to Feb. 10 in China, so *A* and *B* correspond to the conversion rates of Jan. 23 to Feb. 10 respectively. In Eq. (2), *t*_0_ represents the time when the conversion rate is the average of *A* and *B*, and *k* can be directly determined by the rate of change and the derivative value of the conversion rate at time.

The newly established 2019-nCoV modified SEIR propagation dynamics system model is as follows

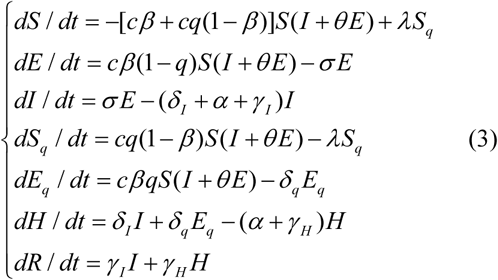

where *δ* _*I*_ is the quarantine rate of the infected, *γ* _*I*_ is the recovery rate of the infected, *δ* _*q*_ is the transformation rate from the exposed to the isolated infected, *γ* _*H*_ is the recovery rate of the isolated infected, is *α* the disease-induced death rate, and *σ* is the transformation rate from the exposed to the infected.

The epidemic data used in this paper comes from the raw epidemic notification data published on the official website of the National Health Commission of the people’s Republic of China (http://www.nhc.gov.cn/). In this paper, the governing Eqs. 2 and 3 is solved using Euler’s numerical method, and the integration step is 0.01 (days). The initial value of the dynamic system refers to the epidemic data officially released by the Chinese government on January 23, 2020, and some parameters have been reasonably estimated. The specific parameter values are shown in Table 1. The other model parameters are the results of fitting optimization based on the original data.

**Table 1.**
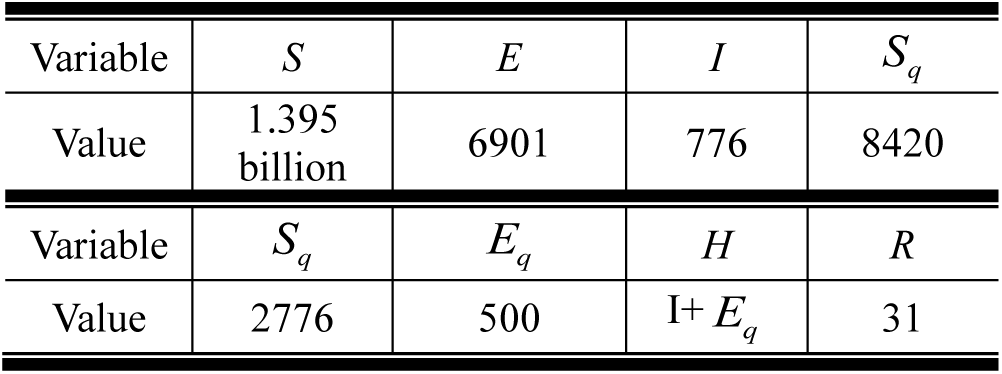
Initial values of the dynamic system

| Variable | $S$ | $E$ | $I$ | $S_q$ |
| --- | --- | --- | --- | --- |
| Value | 1.395 billion | 6901 | 776 | 8420 |
| Variable | $S_q$ | $E_q$ | $H$ | $R$ |
| Value | 2776 | 500 | $I + E_q$ | 31 |

First of all, under different infectious ability of patients with latent period (*θ* =0, 0.6,1), the corresponding model parameters are obtained by fitting and optimization based on the raw data, and predict the epidemic situation in future. As can be seen from the Fig. 2, the corresponding optimal model parameters are found with different infectious abilities. Under the optimal parameters, the number of infected persons predicted by the theoretical model is in good agreement with the raw data from January 23, 2020 to February 10, 2020. Next, we can analyze the effect of different infectious ability of patients in latent period on model estimation. Note that without considering the infectious capacity of patients in the incubation period, that is, *θ*=0, the present model can degenerate to obtain the recent dynamic model of infectious disease transmission established in Ref.[8]. The results shown in Figure 2 show that the peak estimate of the number of infected people using the theory established in Ref.[8] is much higher than the estimate made by the present theoretical model. This is due to the neglect of the infectious ability of patient in latent period in previous models^[6-8]^. Because only the infected are transmissible, the actual probability of infection needs to be overestimated to adequately fit the raw data, which ultimately leads to overestimation of the number of infected people.

**Figure 2.**
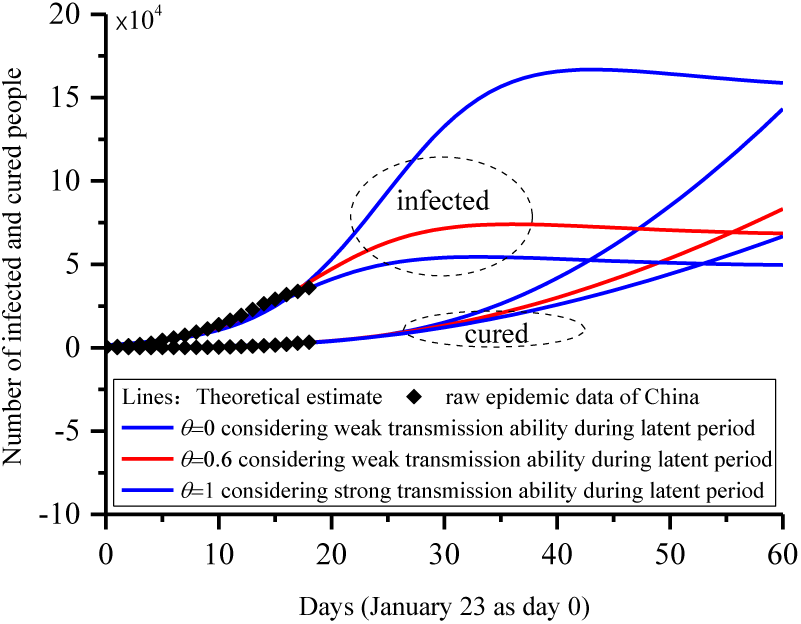
Effect of infectious ability of patient in latent period on the theoretical estimation.

Next, Figure 3 analyzes the effect of variation of incubation period length on the theoretical estimation. It can be seen from Fig. 3 that the peak number of infected persons in the model considering the variation of the incubation period is lower than that in the model with the constant incubation period. This corresponds to an easily understood fact. With the development of the epidemic, the length of the incubation period has shortened, which directly leads to the patients in the incubation period to show symptoms faster and be taken to hospital and quarantined. Therefore, the gradual shortening of the incubation period is obviously conducive to suppressing the spread of the epidemic, and eventually leading to a reduction in the peak number of infections.

**Fig 3.**
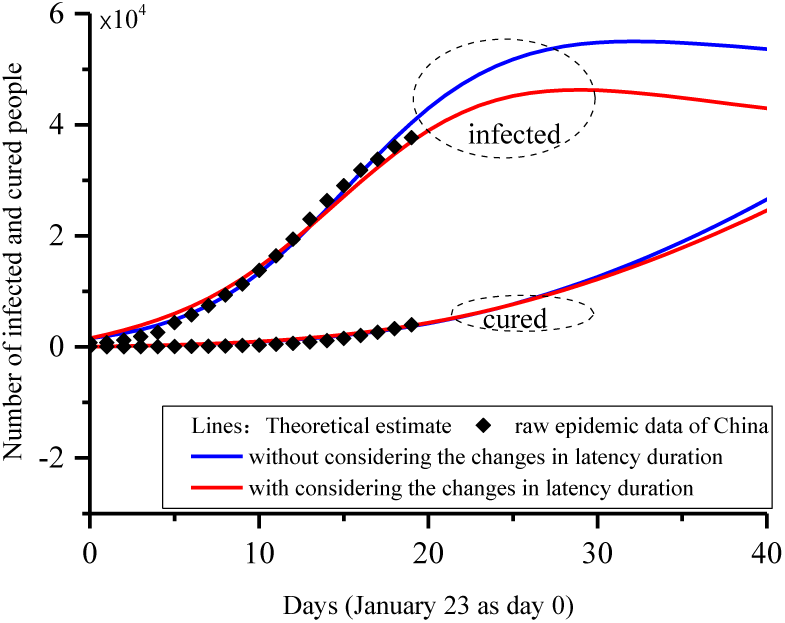
Impact of variation of incubation period length on the theoretical estimation.

**Fig 4.**
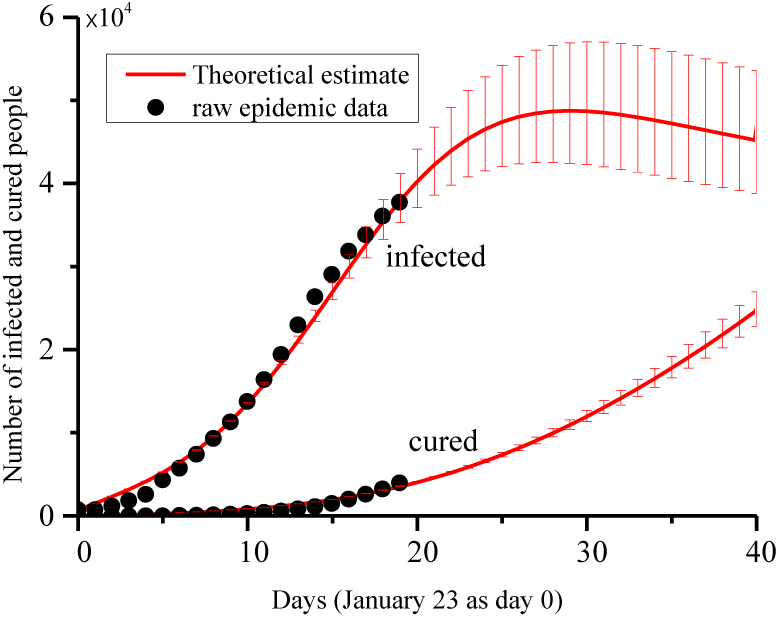
Evaluation of the spread of China’s 2019-nCoV epidemic.

Finally, the trend of China’s 2019-nCoV epidemic was evaluated by using the new present SEIR model. In order to obtain an interval estimate of the peak number of infections and its occurrence time, the contact rate was set to vary within the interval of [2.8, 4.2]. The prediction of the current theoretical model indicates that the number of 2019-nCoV infections in China reaches its peak after February 19, and the upper and lower limits of the estimated peak time of the epidemic respectively correspond to the abscissa of the peaks of the upper and lower envelopes of the theoretical forecast data of the number of infected. The current theoretical estimation of the number of infected people in China is in good agreement with the raw number of infected people in the period from January 23, 2020 to February 10, 2020. It is worth noting that the theoretical model and parameters of this paper were initially established before February 5. The prediction results of the number of infected persons from February 6 to February 10 are mainly consistent with the actual data of subsequent reports, which initially proves the feasibility of the model in predicting the short-term development situation of the epidemic.

This paper attempts to estimate the short-term development of China’s 2019-nCoV epidemic and predicts that the number of people infected in China will peak after February 19. It should be noted that because the infectious ability of patient in latent period and variation of incubation period length are considered, our present model in this paper is closer to the real situation, so the estimates based on it should also be more reliable. However, some necessary assumptions are still made during the model establishment in this paper. The mathematical descriptions made by the model are different from the complex reality, which leads to the inevitable deviation of the prediction results.

This work was financially supported by the National Natural Science Foundation of China (No. 11802225).

## Data Availability

The data used to support the findings of this study are available from the corresponding author upon request.

## Appendix (Matlab code program)

~~~
clc;clear
%%SEIR Transmission dynamics model of 2019 nCoV coronavirus with considering the weak infectious ability and changes in latency duration%%%%%%
%%Shi Pengpeng,Cao Shengli,Feng Peihua,Completed on Jan 5th, first released on Jan 7th%%
%%This latest modified version was released on Feb. 10th, 2020%%
%% Model parameters
c=3.6;beta=6.93e-11;
delta_I=0.13; delta_q=0.13;
gama_I=0.003; gama_H=0.009;
q=9e-7; alpha=0.0001;
theta=0.6; lam=1/14;
T=40; t=0.1; NN=T/t;
%% Initial values
S=13.95e8; E=4007; I=776; Sq=8420;
Eq=3000; H=I+Eq; R=34; De=25;
AA=[S E I Sq Eq H R];
for ii=1:NN
 %% Modified SEIR Transmission dynamics model
 sigma=(1/7-1/3)./(1+exp((ii*t-4)./ii*t/0.2))+1/3;
 dS = -(beta*c+c*q*(1-beta))*S*(I+theta*E)+lam*Sq;
 dE = beta*c*(1-q)*S*(I+theta*E)-sigma*E;
 dI = sigma*E-(delta_I+alpha+gama_I)*I;
 dSq = (1-beta)*c*q*S*(I+theta*E)-lam*Sq;
 dEq = beta*c*q*S*(I+theta*E)-delta_q*Eq;
 dH = delta_I*I+delta_q*Eq-(alpha+gama_H)*H;
 dR = gama_I*I+gama_H*H;
 dDe = alpha*(I+H);
 %% Euler integration algorithm
 S =S+dS*t;
 E = E+dE*t;
 I = I+dI*t;
 Sq = Sq+dSq*t;
 Eq = Eq+dEq*t;
 H = H+dH*t;
 R = R+dR*t;
 AA=[AA; S E I Sq Eq H R];
end
%% Theoretical estimation
Infected(:,1)= round(AA(1:1/t:size(AA,1),3));
Cured(:,1)=round(AA(1:1/t:size(AA,1),7));
plot(0:T,[Infected Cured])
%% raw data
hold on
data_Infected=[776 776 1218 1880 2630 4369 5762 7440
9336 11319 13779 16402 19413 22980
26343 29034 31826 33788 36043 37693]’;
data_Cured=[34 34 38 49 51 60 103 126 171
243 328 475 632 892 1153 1540 2050 2651 3283 3998]’;
plot([1:length(data_Infected)]’-1,[data_Infected data_Cured],’*’)
~~~

